# Seroprevalence of antibodies against Diphtheria, Tetanus and Pertussis over a 12-year period in children in Kilifi, Kenya (2009-2021)

**DOI:** 10.1101/2025.10.07.25337523

**Authors:** CN Mburu, J Ojal, R Selim, R Ombati, D Akech, B Karia, J Tuju, A Sigilai, G Smits, PGM van Gageldonk, FRM van der Klis, S Flasche, EW Kagucia, JAG Scott, IMO Adetifa

**Affiliations:** Epidemiology and Demography Department, KEMRI-Wellcome Trust Research Programme, Kilifi, Kenya; Department of Infectious Diseases Epidemiology, London School of Hygiene and Tropical Medicine, London, United Kingdom; Department of Immunosurveillance, Centre for Infectious Diseases Control, National Institute of Public Health and the Environment (RIVM), The Netherlands; Charite Centre for global Health, Charite-Universitaetsmedizin, Berlin, Germany

## Abstract

**Background:** In Kilifi, no diphtheria or tetanus cases and only sporadic pertussis cases have been reported but the absence of disease does not guarantee sustained immunity. Case surveillance cannot reveal age-specific gaps or waning protection. We conducted serial seroprevalence studies to estimate population immunity trends for diphtheria, pertussis, and tetanus.

**Methods:** We analysed randomly selected participants from multiple cross-sectional surveys within the Kilifi Health and Demographic Surveillance System (KHDSS). IgG antibodies were measured using a fluorescent bead-based multiplex immunoassay applying standard protective thresholds ≥0.011 IU/mL for diphtheria and tetanus. Pertussis antibodies were grouped by likely time since infection. Bayesian multilevel regression with post-stratification was used to adjust estimates for population structure and assay performance and associations with age, sex and year were assessed using logistic regression.

**Results:** Diphtheria seroprotection was low; only 5% of children had long-term seroprotection, with full protection ranging from 11% to 34% and minimal seroprotection from 40% to 52%. Minimal seroprotection increased significantly over time (τ=0.68, p=0.04). Tetanus protection was consistently higher, with long-term seroprotection ranging from 10% to 39% and susceptibility <1% in all years; trends were not significant. Older age was associated with lower seroprevalence for both diphtheria and tetanus. Among adults <1% had long-term diphtheria seroprotection versus 36% for tetanus. Pertussis circulation was minimal with 5% of children and <1% of adults showing antibody concentrations consistent with recent infection.

**Conclusion:** Despite measurable immunity gaps, particularly for diphtheria, no outbreaks of diphtheria or tetanus have been observed in Kilifi over the past decade, and tetanus protection remains high. These findings suggest that the current immunisation programme continues to provide effective population level protection. Serology nonetheless highlights vulnerable cohorts and provides a baseline for monitoring susceptibility over time. Sustaining high coverage and maintaining sensitive surveillance will be essential to detect any future risk early and guide decisions on booster doses if susceptibility increases.

## Background

Routine childhood immunisation schedule in Kenya was formalised in 1980 with the introduction of Kenya Expanded Programme of Immunisation (KEPI) (1). The Diphtheria-Pertussis-Tetanus (DPT) schedule initially consisted of three doses of a trivalent vaccine at 6,10 and 14 weeks. Presently, the schedule includes three doses of DPT as a pentavalent vaccine combined with *Haemophilus influenzae* type B (hib) and Hepatitis B vaccines since November 2001 (2). Although WHO recommends 3 booster doses for DPT to be administered at 12-23 months, 4-7 years and 9-15 years of age to ensure long term protection, Kenya is yet to implement any of these (3,4).

Kenya has made significant progress towards Vaccine Preventable Diseases (VPDs) elimination. For instance, maternal and neonatal tetanus were verified to be eliminated in 2019 (5) and reported pertussis cases have declined though surveillance gaps limit interpretation (6). Between 2009 and 2018, national pentavalent coverage ranged from 75–81%, below the 90% target set by the Global Vaccine Action Plan (7,8). In addition, coverage heterogeneity across regions may leave some subpopulations vulnerable and surveillance alone cannot reveal age-specific immunity gaps or waning protection.

Serosurveillance provides a complementary approach by directly quantifying antibody prevalence and identifying susceptible cohorts. Historical experience underscores this value. In the United Kingdom, Hib disease re-emerged within a decade of vaccine introduction due to waning immunity and use of a less immunogenic vaccine formulation with subsequent serological studies confirming low antibody concentrations in affected cohorts and guiding catch-up and booster programmes (9,10). In the former Soviet Union, serosurveys revealed widespread adult susceptibility to diphtheria preceding a large epidemic (11) and across Europe, measles serosurveys identified inadequate measles immunity in several countries despite low reported incidence (12) predicting the large outbreaks that followed (13).

This study describes age-specific seroprevalence of diphtheria, pertussis, and tetanus among children and adults in Kilifi, providing population-level immunity profiles, identifying vulnerable cohorts, and underscoring the importance of sustaining high coverage in the routine immunisation programme.

## Methods

### Survey design and study population

Our study, previously described (14), used a retrospective series of cross-sectional serum samples from the Kilifi Health & Demographic Surveillance System (KHDSS) population, sourced from three previous serological studies in the KEMRI-Wellcome Trust Research Program (KWTRP) biobank. These observational studies included residents registered in KHDSS, which tracks births, deaths, and migration within a population of over 300,000 people in Kilifi County, home to approximately 1.4 million residents (15).

Plasma or serum samples collected between 2009 and 2019 were obtained from three survey frameworks: (i) the Malaria Cross-Sectional Survey (2009–2013) (16) conducted annually as an independent, random cross-sectional sample nested within a rolling cohort of healthy children in the KHDSS, this cohort has continuously recruited children since 1998, following them until age 15; (ii) the Pneumococcal Conjugate Vaccine Impact Study (PCVIS) (2015–2019)(17) which conducted annual cross-sectional surveys of KHDSS residents; and (iii) the COVID-19 serosurveillance study conducted in 2021 in response to the pandemic, randomly sampling KHDSS residents (18). Sera from adults (≥15 years) were only available from the 2021 study.

To examine the years 2009 to 2013, we took an age-stratified random sub-sample of the Malaria Cross-Sectional Surveys conducted in 2009, 2011, and 2013; the age strata were 0, 1, 2, 3, 4, 5, 6, 7, 8–9, and 10–14 years and sera from 50 children were sampled per stratum (19). We analysed sera from all of the participants in the PCVIS study (2015–2019) and the COVID-19 serosurveillance study (2021). In the PCVIS study, participants were selected using an independent, age-stratified random sampling method from the KHDSS population, with 50 children randomly chosen per stratum across 10 age groups (0, 1, 2, 3, 4, 5, 6, 7, 8–9, and 10–14 years). For the 2021 study, 100 individuals were randomly selected for each 5-year age band from 0–14 years, 50 individuals for each 5-year age band from 15–64 years, and 50 individuals aged ≥65 years, totalling 850 participants.

In the original studies, approximately 2 mls of venous blood was collected from all consenting participants, then separated, aliquoted, and stored at -70°C until processing.

### Bead coupling and fluorescent bead multiplex immunoassay

We coupled five antigens to fluorescent beads: Pertussis toxin (Ptx), Filamentous hemagglutinin (FHA), and Pertactin (Prn) from *Bordetella pertussis*, and diphtheria and tetanus toxins. The immunoassay was performed as previously described (20). Briefly, serum/plasma samples (25 µL) were diluted in duplicate (1/200 and 1/4000) in an assay buffer containing phosphate-buffered saline, 0.1% Tween 20, and 3% bovine serum albumin. For high-concentrations sera, an additional 1/16,000 dilution was included to ensure measurements fell within the linear range of the assay.

The in-house Diphtheria-Tetanus (DT) luminex standard was the same reference described by Gageldonk et al (20) obtained from a DTP-vaccinated adult, with serum values assigned in IU/ml using the Toxin Binding Inhibition (ToBI) assay for both toxins. In-house references used in ToBI were calibrated against international standards (NIBSC codes Di-03 and TE-03). The in-house pertussis standard was derived from a pool of 28 diagnostic sera submitted to RIVM for suspected whooping cough, confirmed by high anti-Ptx IgG concentrations. These were historically calibrated against the U.S. Reference Pertussis Antiserum Human Lot 3 (for Ptx IgG and FHA IgG) and Lot 4 (for Prn IgG) from CBER/FDA, with results expressed in ELISA Units (EU/mL). This calibration was maintained to ensure comparability with earlier studies(20), though subsequent RIVM assays have been re-standardised to the 1st International Standard 06/140 expressed in IU/mL(21,22)

To monitor assay performance, high and low in-house serum controls, standards and blanks were included on each plate. Analyte-specific QC trend analysis was conducted for every plate assayed and the linearity and recovery of the standards were checked to confirm they were within the acceptable range (80%-120%). Samples were also assayed in dependent duplicate dilutions (1/200 and 1/4000), and the coefficients of variation (CVs) of the IgG concentrations were checked to ensure they were within the acceptable range (20).

For each analyte, median fluorescent intensity (MFI) values were converted to IU/mL (for diphtheria and tetanus) or EU/mL (for pertussis) by interpolation from a five-parameter logistic (log–log) standard curve. Results were determined from the mean of all dilutions that fell within the linear range of the standard curve.

The Correlates of Protection (CoPs) for both diphtheria and tetanus have been extensively studied from observational data and vaccine efficacy trials. Serum IgG concentrations of approximately 0.01–0.02 IU/mL are considered the minimum level associated with short-term protection (23) while concentrations ≥0.1 IU/mL are widely accepted as indicative of clinical protection against both diseases (3,4). Higher concentrations (≥1.0 IU/mL) do not imply lifelong immunity but are associated with longer persistence of protective levels and thus are often used as a target for durable protection (24). Because both diphtheria and tetanus are toxin-mediated diseases, these thresholds are applied to both although antibody waning is typically more pronounced for diphtheria than for tetanus. Based on these definitions’ participants with anti-tetanus or anti-diphtheria IgG ≥0.011 IU/ml were considered protected and further stratified into minimal (0.011≤IgG<0.1IU/ml), full (0.1≤IgG<1 IU/ml) and long-term (IgG≥1 IU/ml) protection categories.

No protective cut-off was used for pertussis as there are no defined CoPs. Instead, following longitudinal studies in convalescent pertussis cases, which showed how anti-PT IgG levels decline predictably after infection (25–27) antibody concentrations were categorised into four groups reflecting time since infection: ≥125 EU/mL (infection within the past 6 months), 62.5 to <125 EU/mL (within the past 12 months), 20 to <62.5 EU/mL (infection more than 12 months previously and/or vaccination response), and <20 EU/mL (no evidence of recent infection)

### Ethical considerations

Specific ethical approval for this study was obtained from the Scientific and Ethics Review Unit (SERU) of the Kenya Medical Research Institute (Protocol SERU 3847). The serological samples were collected under SERU-approved protocols with a specific consent for storage of residual samples and use in future research (SERU 1433, 4085, 2887, 3149, 3426). Written informed consent was obtained from parents/legal guardians of all participants before sample collection.

### Statistical analysis

We first tabulated the seropositive results in each survey year based on the age strata at the time of the sample collection, sex, ethnic group and location in KHDSS and generated seroprevalence in these age groups.

To analyse changes in seroprevalence over time among children under 15 yrs of age we collapsed the age strata into four: <1 year, 1-4 years, 5-9 years and 10-14 years. To analyse the data for adults above 15 years of age we classified the data using the 5-year age bands defined in the 2021 survey.

During the study period (2009–2019), the age-specific seroprevalence remained relatively stable within individual-year age groups across the four defined age strata. Consequently, we did not calculate a summary weighted seroprevalence while grouping the data into the four age strata for analysis. To enable valid comparisons across years while minimising the influence of differences in age, we applied direct age standardisation. We used the average age distribution of the KHDSS population across the study period (2009–2019) as the reference population, and standardised both age-specific and overall seroprevalence estimates to this structure. This standardisation was implemented using multilevel regression and post-stratification (MLRP) which also adjusted for sensitivity and specificity of the assay. The sensitivity was estimated at 99% for all five antigens with specificities of 92% for diphtheria,93% for tetanus and 94% for pertussis antigens (Ptx, FHA, Prn) based on validation studies comparing Luminex results to toxin-binding inhibition assays (diphtheria, tetanus) or FDA/CBER reference sera (pertussis). These estimates were derived from European reference populations and represent the best available validation for this platform (20). The MLRP was implemented by fitting a Bayesian logistic regression that included age as a variable (28). Non-informative priors were used for all the parameters and models were fitted using rjags software package. Bayesian population-weighted and test-adjusted seroprevalence estimates were visualized using bar graphs, and group differences within each year were assessed using chi-square or Fisher’s exact tests, as appropriate. A multivariate logistic regression model was used to examine associations between seroprevalence and year, sex, age, location, and ethnic group.

Geometric mean concentrations (GMCs) and the 95% confidence levels were also calculated, and values tabulated after adjusting for underlying population. To display the variability in age-specific IgG concentrations, raw antibody concentrations were log transformed and displayed via boxplot for each survey year. One-way ANOVA was used to test differences in GMCs between groups in each year. A Mann-Kendall trend test was employed to assess whether there was a statistically significant trend in seroprevalence and GMCs across the survey years. Statistical analyses were conducted using the R-statistical software (29).

## Results

The study population comprised of 2686 participants of which 1381(51%) were males. The median age of the participants was 6 yrs. (IQR: 3-8 yrs.). Ages were comparable between males (median:5 IQR:3-8) and females (median: 6, IQR:3-8, p=0.198). The number of samples assayed each year varied from 290 to 520. Estimates of diphtheria seroprotection were much lower than those for tetanus seroprotection and they declined with age. The majority of participants were from Chonyi and Giriama ethnic groups (Table 1).

**Table 1.**
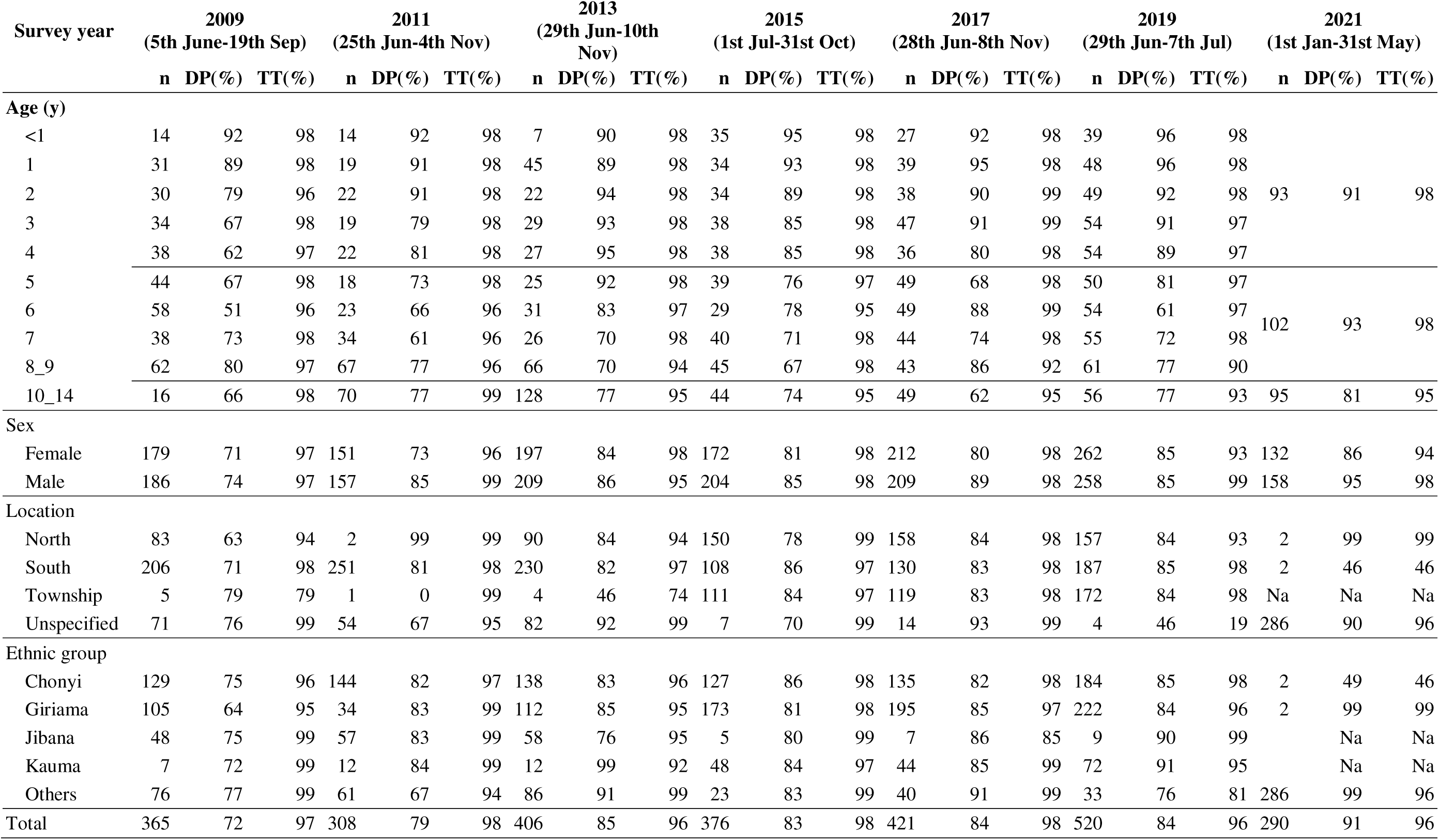
Crude proportion of participants with minimal seroprotection ((≥0.011IU/ml) against tetanus (TT) and diphtheria (DP) and total number of participants per survey year (n) stratified by age strata during sample collection, sex, location in KHDSS and ethnic group.

### Immunity against Diphtheria

The majority of children had minimal seroprotection against diphtheria in all the survey years which ranged between 40% CI: (34-46%) in 2011 and 52% CI: (45-58%) in 2013 (Table s1 and Fig.1). The proportion of children with full seroprotection ranged between 11% CI: (05-22%) in 2009 and 34% CI: (26-43%) in 2021. Less than 5% of children had long-term seroprotection against diphtheria in every year. Significant heterogeneity in seroprevalence levels with minimal seroprotection across ages was observed in all surveys (P<0.05). A Mann-Kendall trend test indicated a significant increasing trend in diphtheria minimal seroprotection across the survey years(tau=0.68, P=0.04).

**Figure 1.**
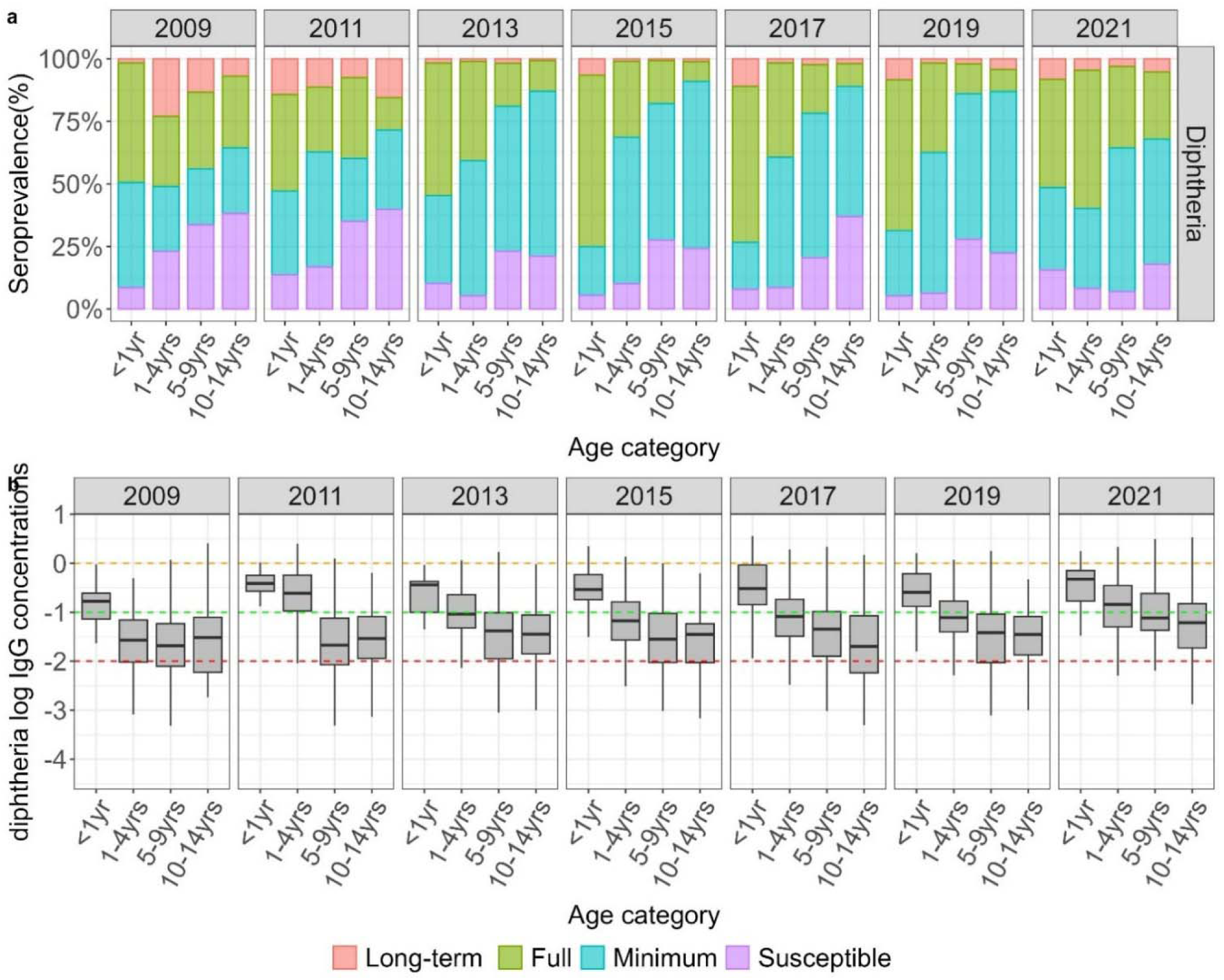
a. Standardized age-specific diphtheria seroprevalence estimates in children by calendar year adjusted for test performance using Bayesian multilevel regression and poststratification. b. Distribution of log IgG concentration showing the median and IQR. The red dotted line is the threshold for minimal seroprotection (0.011≤IgG<0.1 IU/ml), the green dotted line is the threshold for full seroprotection (0.1≤IgG<1 IU/ml) and the orange dotted line is the threshold for long-term protection (IgG≥1 IU/ml).

A comparable pattern was observed in GMC levels, showing significant variation across age groups (P <0.05), a decline with age (Table s2 and Fig.1) and an increasing trend across the years (tau=0.411) although this was not significant. GMC levels in children under one year exceeded the full protection threshold in majority of the surveys, while those in children aged one to nine were below the full seroprotection threshold but above the minimum seroprotection threshold. In some years, GMCs in the oldest age group dropped below the minimum seroprotection threshold (Fig.1).

In the multivariate logistic regression, older children had significantly lower seroprevalence than infants, males had significantly higher seroprevalence compared to females (aOR=1.41,95%CI:1.15-1.75) and all subsequent years demonstrated significantly higher seroprevalence than the baseline year 2009 (Table 2).

Among adults (≥15 years), although seroprevalence was >68%, long-term seroprotection against diphtheria was found in <1% (Table s3 and Fig.S1).

**Table 2.** Risk factors associated with seroprevalence of diphtheria, tetanus, and pertussis.

| **Predictors** | **Tetanus**<br>OR (95% CI) | **Diphtheria**<br>OR (95% CI) | **Pertussis**<br>OR (95% CI) |
| --- | --- | --- | --- |
| **Sex** | 1 | 1 | 1 |
| **Male** | 1.39 [0.93-2.10] | 1.41 [1.15-1.75] | 1.27 [1.23-3.13] |
| **Age category** |  |  |  |
| **<1yr** | 1 | 1 | 1 |
| **1-4yrs** | 0.04 [0.02-0.05] | 0.12 [0.02-0.40] | 0.96 [0.65-1.40] |
| **5-9yrs** | 0.01 [0.00-0.02] | 0.05 [0.01-0.15] | 1.12 [0.76-1.63] |
| **10-14yrs** | 0.01 [0.00-0.06] | 0.04 [0.01-0.12] | 0.72 [0.47-1.07] |
| **Survey year** |  |  |  |
| **2009** | 1 | 1 | 1 |
| **2011** | 1.34 [0.55-3.36] | 1.61 [1.11-2.36] | 0.92 [0.66-1.30] |
| **2013** | 1.12 [0.51-2.40] | 2.54 [1.75-3.69] | 0.95 [0.69-1.30] |
| **2015** | 1.39 [0.58-3.38] | 1.78 [1.21-2.64] | 0.98 [0.71-1.38] |
| **2017** | 1.54 [0.66-3.65] | 2.11 [1.44-3.11] | 1.16 [0.84-1.61] |
| **2019** | 0.81 [0.37-1.67] | 2.07 [1.43-3.01] | 1.01 [0.74-1.39] |
| **2021** | 0.87 [0.31-2.35] | 3.72 [2.17-6.47] | 0.63 [0.42-0.93] |
| **Ethnic group** |  |  |  |
| **Chonyi** | 1 | 1 | 1 |
| **Jibana** | 1.57 [0.59-5.41] | 0.87 [0.58-1.33] | 1.25 [0.88-1.82] |
| **Giriama** | 1.18 [0.62-2.22] | 0.98 [0.71-1.35] | 0.85 [0.65-1.11] |
| **Kauma** | 1.02 [0.41-2.84] | 1.37 [0.83-2.33] | 1.01 [0.68-1.50] |
| **Other** | 0.71 [0.26-2.57] | 0.76 [0.42-1.43] | 0.65 [0.40-1.05] |
| **Location** |  |  |  |
| **South** | 1 | 1 | 1 |
| **North** | 0.58 [0.29-1.13] | 0.87 [0.62-1.24] | 1.14 [0.86-1.51] |
| **Township** | 0.87 [0.38-2.04] | 0.87 [0.58-1.32] | 1.13 [0.81-1.57] |
| **Unspecified** | 1.57 [0.37-5.77] | 1.40 [0.69-2.75] | 1.62 [0.93-2.78] |

To define prevalence, we used the basic seroprotection threshold for tetanus and diphtheria (>0.011 IU/mL) and for pertussis we used the threshold of recent infection (>62 EU/mL). The analysis was based on the raw estimates shown in Table 1. Explanatory factors included survey year (with 2009 as the reference), age as a categorical variable (<1 year as the reference), sex (female as the reference), location (south as the reference), and ethnic group (Chonyi as the reference). Location and ethnic group were not significantly associated with tetanus, diphtheria or pertussis seroprevalence

### Immunity against Tetanus

Significant heterogeneity in seroprevalence levels across ages was observed in most of the years (P<0.05) and in most calendar years, the strength of seroprotection to tetanus declined with increasing age (Table 2,Table S1, Fig. 2). A Mann-Kendall trend test indicated a decreasing trend in tetanus minimal seroprotection across the survey years, but this was not significant (tau=-0.28). Among infants, majority had long-term seroprotection, among those aged 1-9 years the majority had full seroprotection and among the oldest age group the majority had minimal seroprotection. The proportion of children susceptible to tetanus was consistently less than 1% in all surveyed years (Table s1 and Fig.2).

**Figure 2.**
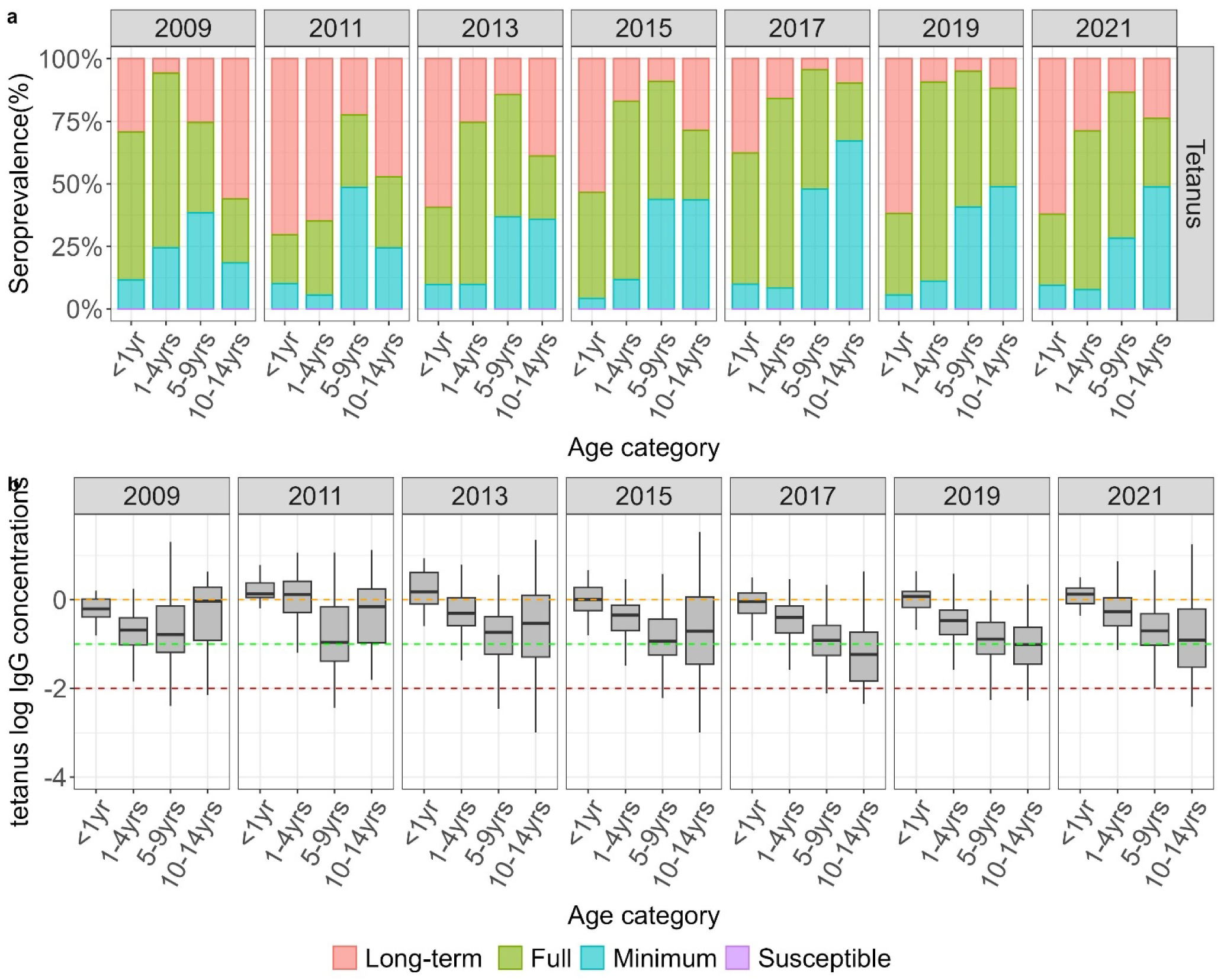
a. Standardized age-specific tetanus seroprevalence estimates by calendar year adjusted for test performance using Bayesian multilevel regression and poststratification. b. Distribution of log IgG concentrations with median and IQR. The red dotted line is the threshold for minimal seroprotection (0.011≤IgG<0.1 IU/ml), the green dotted line is the threshold for full seroprotection (0.1≤IgG<1 IU/ml) and the orange dotted line is the threshold for long-term protection (IgG≥1 IU/ml).

While GMC levels exhibited significant variation across age groups (P <0.05) and decreased with age (Table s2 and Fig.2), the waning observed was less pronounced compared to diphtheria. Log IgG concentrations across all age groups remained above the full seroprotection thresholds, except for the oldest age group, where the concentrations fell below the full seroprotection threshold in certain years (Fig.2).There was a declining trend in GMCs across the survey years, but this was also not significant(tau=-0.19).

In the multivariate logistic regression, older children had significantly lower seroprevalence than infants. Tetanus seroprevalence did not vary significantly with sex or calendar year (Table 2). The proportion of adults with long-term seroprotection against tetanus was 36%. Adult females had slightly higher seroprotection levels compared to adult males (Fig s2 and Table s3).

### Likely time of infection for Pertussis

The majority of children in Kilifi have not recently experienced a pertussis infection, evident from the high proportion of IgG levels between 0-20 EU/ml across all age groups in all survey years (Table s4 and Fig. 3). This proportion varied from 49% CI: (42-55%) in 2021 to 63% CI: (34-73%) in 2017. About 5% of children, across different age groups, exhibited IgG concentrations compatible with pertussis infection in the past 12 months (IgG≥62.5 IU/ml) in all survey years. The proportion of children with either a vaccination response and/or infection occurring >12 months ago ranged from 11% CI: (03-21%) in 2009 to 21% CI: (16-27%) in 2021.

**Figure 3.**
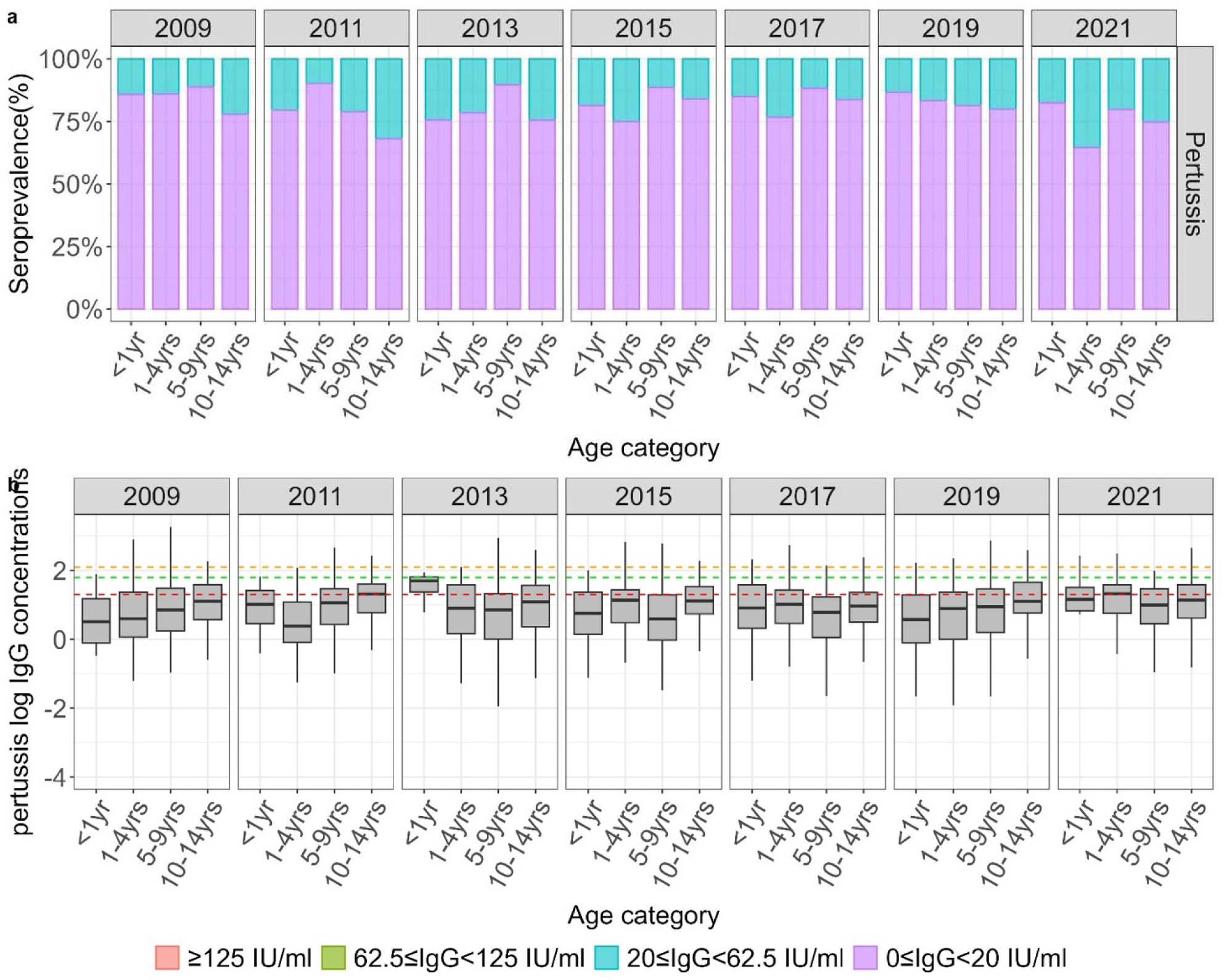
a. Standardized age-specific pertussis seroprevalence estimates by calendar year adjusted for test performance using Bayesian multilevel regression and poststratification. b. Distribution of log IgG concentrations with median and IQR. The orange dotted line is the threshold for IgG levels equal or above 125 EU/mL (infection in the past 6 months), the green dotted line is the threshold for IgG levels equal or above 62.5 (infection in the past 12 months) while the red line is the threshold for IgG levels equal or above 20 EU/mL (infection in the past>12 months or vaccination response). Log IgG levels below the red line are below 20 EU/mL (no recent infection).

There was a significant heterogeneity in GMC levels across the ages in some of the years where levels decreased with increasing age up to around 4 years and then increased afterwards but this trend was not consistent across the years (Table s4 and Fig.3). Among adults, 63% (CI: 58-67%) have IgG concentrations suggesting they have not experienced a recent infection while 23% (CI: 19-27%) had concentrations suggesting an infection >12 months ago (Table s5 and Fig.s3).

In the logistic regression analysis, males were more likely to not have had a recent pertussis infection compared to females (aOR=1.27, 95% CI:1.23-3.13). Neither age, sex nor calendar year were associated with an antibody concentration above 62.5 EU/ml which is suggestive of infection in the last year (Table 2).

## Discussion

This study provides the first population based serological profile of diphtheria, tetanus and pertussis among children in Kilifi with an additional snapshot in adults. We found inadequate diphtheria immunity: beyond infancy, most children had IgG levels below full protection in all survey years, well under the 84–89% herd immunity threshold (30,31). Among adults, only 20% achieved full seroprotection and just 1% maintained long-term seroprotection. By contrast, tetanus seroprevalence was consistently high with 70-80% of children fully protected across the study period though some older children had minimal protection. For pertussis, only 5% of children and less than 1% of adults had antibody concentrations ≥62.5 EU/ml, suggestive of an infection in the last 12 months.

To interpret these findings, it is important to triangulate them with local coverage and disease surveillance data. KHDSS data from 2010–2017 showed an increase in coverage for the three-dose pentavalent primary series from 36% to 81% (32). In our analysis, seroprevalence in infants (<1 year) for both diphtheria and tetanus was at least as high as, and in some years higher than, these coverage estimates which suggests that the primary series is immunogenic and performing as expected in this age group. Slightly higher seroprevalence compared to coverage is plausible, as administrative coverage estimates are susceptible to inaccuracies in dose recording and target population denominators (33). For pertussis, however, the pattern differs: even in infants, the proportion with antibody concentrations consistent with recent vaccination was lower than expected from coverage data. Given that all three antigens are administered in the same shot, this discrepancy most likely reflects limitations in the pertussis correlates of protection rather than programme failure (34,35).

In parallel, over three decades of continuous paediatric surveillance at Kilifi County Hospital have recorded no cases of diphtheria and only sporadic cases of tetanus and pertussis (KHDSS records, unpublished). National data show a similar pattern: Kenya reported no diphtheria cases from 2001 to 2023, with only 27 cases notified in 2024 (36,37) and just 43 pertussis cases nationwide between 2007 and 2022 (38). These observations suggest that clinical disease has been very rare despite the immunity gaps identified in our serology data. This striking absence of diphtheria disease may indicate that the antibody thresholds we applied underestimate true functional immunity in older children and adults or that herd immunity in younger recently vaccinated cohorts has been sufficient to prevent transmission. The findings for tetanus are consistent with both high childhood seroprotection and Kenya’s verification of maternal and neonatal tetanus elimination in 2019 (5). It is also important to recognise that disease surveillance, while robust at KHDSS may still miss mild or community-managed cases, meaning that very low-level transmission could persist undetected. Taken together, these observations support viewing serology as a complementary tool that can highlight emerging immunity gaps, but which must be interpreted alongside coverage and clinical data rather than used in isolation.

Although diphtheria and tetanus are delivered together in the pentavalent vaccine, their immunity profiles differ markedly, a finding also reported among children in Tajikistan (39) and the US (40). Diphtheria antibody concentrations decline rapidly after primary immunisation, with studies showing that up to 67% of children lack protective levels within 3–13 years and concentrations can fall five-fold within the first year (41,42). In contrast, tetanus antibody concentrations decline much more slowly, with estimated half-lives of 11–14 years (43) and the toxoid is one of the most immunogenic and thermostable vaccine antigens (44). These differences explain why tetanus seroprotection remained consistently high in our study whereas diphtheria displayed substantial immunity gaps in both children and adults. The implications also vary. For tetanus, herd protection does not apply. Control depends on sustained individual immunity, which in Kenya has been strengthened by routine immunisation and the targeted TTCV programme for women of reproductive age (45), culminating in verification of maternal and neonatal tetanus elimination in 2019 (5). For diphtheria, herd protection is more complex: the toxoid vaccine prevents toxin-mediated disease but does not fully block colonisation or carriage of *Corynebacterium diphtheriae* (46). Nonetheless, by reducing symptomatic cases and shortening carriage duration, high childhood coverage reduces the force of infection and suppresses organism circulation as seen in Western Europe where outbreaks have remained rare despite low adult antibody concentrations (46–48).

Pertussis presented a different picture from diphtheria and tetanus. Most children had antibody concentrations below 20 EU/ml and only around 5% had concentrations consistent with infection within the past year suggesting low ongoing circulation. Similar findings have been reported from The Gambia and Senegal (27,49), where a small but detectable proportion of children showed evidence of recent infection. Interpretation of these findings is challenging as there are no universally agreed CoPs for pertussis (25,50), antibody concentrations wane rapidly (51,52) and distinguishing vaccine-induced from infection-induced antibodies is difficult (53). We measured responses to Ptx, FHA, and Prn antigens but did not attempt to classify individuals as recently infected versus vaccinated, as these relationships remain incompletely understood. Consequently, the apparent discrepancy between high coverage and relatively low antibody concentrations may reflect limitations in serological interpretation rather than programme weakness.

A key strength of this study is the use of multiple, randomly sampled cross-sectional surveys spanning a decade, providing a rare temporal perspective on population immunity in a well-defined population. The large sample size and the use of a validated multiplex bead-based immunoassay added precision and allowed simultaneous, standardised measurement of antibodies to multiple antigens across years. Combined with detailed local coverage data this dataset enabled an unusually granular triangulation of serology and vaccination coverage that is rarely possible in LMIC settings.

Several limitations should also be acknowledged. Adult data were available only from 2021, constraining inferences about longer-term trends in adult immunity. CoPs are best established in infants and low antibody concentrations in older children and adults may not fully reflect susceptibility, as memory responses can remain protective (50). In addition, assay performance estimates were derived from European reference populations, which, despite calibration to international standards, may not perfectly capture immune responses in Kilifi (20). Finally, disease surveillance and antibody data do not always align perfectly; the absence of diphtheria cases despite measurable serological susceptibility highlights the need to interpret antibody data as complementary rather than definitive.

Our results show that tetanus protection remains consistently high, aligning with the absence of paediatric cases and Kenya’s achievement of maternal and neonatal tetanus elimination. In contrast, diphtheria seroprotection was suboptimal in both children and adults, yet no diphtheria cases have been observed in over three decades of hospital surveillance, suggesting that current coverage is still sufficient to prevent transmission or that functional immunity persists despite low antibody concentrations. Pertussis circulation appears minimal, consistent with clinical data showing few reported cases, though serology cannot easily distinguish recent infection from waning vaccine responses. Taken together, these findings indicate that the Kenyan immunisation programme continues to provide effective public health protection against all three diseases. The primary implication is a need to sustain high coverage and continue serological and clinical surveillance to detect emerging susceptibility early and inform future decisions about boosters or catch-up interventions if the risk increases.

## Data Availability

All data produced in the present study are available upon reasonable request to the authors

## References

1. Cines C, Immunizatio N. National policy guidelines on immunization 2013. Nairobi Minist Health [Internet]. 2014 [cited 2025 May 27]; Available from: https://devinit.org/files/media/immunization_policy_guidline.pdf

2. Ndiritu M, Cowgill KD, Ismail A, Chiphatsi S, Kamau T, Fegan G, et al. Immunization coverage and risk factors for failure to immunize within the Expanded Programme on Immunization in Kenya after introduction of new Haemophilus influenzae type b and hepatitis b virus antigens. BMC Public Health. 2006 May 17;6(1):132.

3. Tetanus vaccines: WHO position paper, February 2017 – Recommendations. Vaccine. 2018 June;36(25):3573–5.

4. World Health Organization. Diphtheria vaccine: WHO position paper, August 2017 – Recommendations. Vaccine. 2018 Jan;36(2):199–201.

5. Kenya Attains Maternal and Neonatal Tetanus Elimination | Science Africa [Internet]. [cited 2025 Sept 8]. Available from: https://news.scienceafrica.co.ke/kenya-attains-maternal-and-neonatal-tetanus-elimination/

6. WHO. WHO vaccine-preventable diseases: monitoring system. 2020 global summary. 2020.

7. Kenya | Gavi, the Vaccine Alliance [Internet]. [cited 2025 Sept 8]. Available from: https://www.gavi.org/programmes-impact/country-hub/africa/kenya

8. Global vaccine action plan 2011-2020 [Internet]. [cited 2025 Sept 8]. Available from: https://www.who.int/publications/i/item/global-vaccine-action-plan-2011-2020

9. Ladhani S, Ramsay M, Flood J, Campbell H, Slack M, Pebody R, et al. Haemophilus influenzae serotype B (Hib) seroprevalence in England and Wales in 2009. Euro Surveill Bull Eur Sur Mal Transm Eur Commun Dis Bull. 2012 Nov 15;17(46):20313.

10. Cutts FT, Hanson M. Seroepidemiology: an underused tool for designing and monitoring vaccination programmes in low-and middle-income countries. Trop Med Int Health. 2016;21(9):1086–98.

11. Markina SS, Maksimova NM, Vitek CR, Bogatyreva EY, Monisov AA. Diphtheria in the Russian Federation in the 1990s. J Infect Dis. 2000 Feb;181(s1):S27–34.

12. Andrews N. Towards elimination: measles susceptivility in Australia and 17 European countries. Bull World Health Organ. 2008 Mar 1;86(3):197–204.

13. Hens N, Abrams S, Santermans E, Theeten H, Goeyvaerts N, Lernout T, et al. Assessing the risk of measles resurgence in a highly vaccinated population: Belgium anno 2013. Eurosurveillance [Internet]. 2015 Jan 8 [cited 2025 Sept 10];20(1). Available from: https://www.eurosurveillance.org/content/10.2807/1560-7917.ES2015.20.1.20998

14. Mburu CN, Ojal J, Selim R, Ombati R, Akech D, Karia B, et al. Seroprevalence of Immunoglobulin G against measles and rubella over a 12-year period (2009–2021) in Kilifi, Kenya and the impact of the Measles-Rubella (MR) vaccine campaign of 2016. Vaccine. 2025 Aug 13;61:127425.

15. Scott JAG, Bauni E, Moisi JC, Ojal J, Gatakaa H, Nyundo C, et al. Profile: the Kilifi health and demographic surveillance system (KHDSS). Int J Epidemiol. 2012;41(3):650–7.

16. Mwangi TW, Ross A, Snow RW, Marsh K. Case definitions of clinical malaria under different transmission conditions in Kilifi District, Kenya. J Infect Dis. 2005 June 1;191(11):1932–9.

17. PCVIS|Kemri|Wellcome Trust [Internet]. Available from: https://kemri-wellcome.org/programme/pcvis-2/

18. Etyang AO, Adetifa I, Omore R, Misore T, Ziraba AK, Ng’oda MA, et al. SARS-CoV-2 seroprevalence in three Kenyan health and demographic surveillance sites, December 2020-May 2021. PLOS Glob Public Health. 2022 Aug 18;2(8):e0000883.

19. Hammitt LL, Crane RJ, Karani A, Mutuku A, Morpeth SC, Burbidge P, et al. Effect of Haemophilus influenzae type b vaccination without a booster dose on invasive H influenzae type b disease, nasopharyngeal carriage, and population immunity in Kilifi, Kenya: a 15-year regional surveillance study. Lancet Glob Health. 2016 Mar 1;4(3):e185–94.

20. van Gageldonk PG, van Schaijk FG, van der Klis FR, Berbers GA. Development and validation of a multiplex immunoassay for the simultaneous determination of serum antibodies to Bordetella pertussis, diphtheria and tetanus. J Immunol Methods. 2008;335(1–2):79–89.

21. Versteegen P, Pinto MV, Barkoff AM, Gageldonk PGM van, Kassteele J van de, Houten MA van, et al. Responses to an acellular pertussis booster vaccination in children, adolescents, and young and older adults: A collaborative study in Finland, the Netherlands, and the United Kingdom. eBioMedicine [Internet]. 2021 Mar 1 [cited 2025 Oct 6];65. Available from: https://www.thelancet.com/journals/ebiom/article/PIIS2352-3964(21)00040-2/fulltext

22. van der Lee S, Stoof SP, van Ravenhorst MB, van Gageldonk PGM, van der Maas NAT, Sanders EAM, et al. Enhanced Bordetella pertussis acquisition rate in adolescents during the 2012 epidemic in the Netherlands and evidence for prolonged antibody persistence after infection. Eurosurveillance. 2017 Nov 23;22(47):17–00011.

23. Ipsen J. Circulating antitoxin at the onset of diphtheria in 425 patients. J Immunol. 1946;54(4):325–47.

24. The Immunological Basis for Immunization Series Module 3: Tetanus [Internet]. [cited 2025 Sept 4]. Available from: https://www.who.int/publications/i/item/9789241513616

25. De Greeff SC, De Melker HE, Van Gageldonk PGM, Schellekens JFP, Van Der Klis FRM, Mollema L, et al. Seroprevalence of Pertussis in the Netherlands: Evidence for Increased Circulation of Bordetella pertussis. Ratner AJ, editor. PLoS ONE. 2010 Dec 1;5(12):e14183.

26. De Melker HE, Versteegh FGA, Conyn-van Spaendonck MAE, Elvers LH, Berbers GAM, Van Der Zee A, et al. Specificity and Sensitivity of High Levels of Immunoglobulin G Antibodies against Pertussis Toxin in a Single Serum Sample for Diagnosis of Infection with Bordetella pertussis. J Clin Microbiol. 2000 Feb;38(2):800–6.

27. Scott S, Van Der Sande M, Faye-Joof T, Mendy M, Sanneh B, Barry Jallow F, et al. Seroprevalence of Pertussis in The Gambia: Evidence for Continued Circulation of Bordetella pertussis Despite High Vaccination Rates. Pediatr Infect Dis J. 2015 Apr;34(4):333–8.

28. Uyoga S, Adetifa IMO, Karanja HK, Nyagwange J, Tuju J, Wanjiku P, et al. Seroprevalence of anti-SARS-CoV-2 IgG antibodies in Kenyan blood donors. Science. 2021 Jan 1;371(6524):79–82.

29. Team RC. R: A language and environment for statistical computing. R Foundation for Statistical Computing, Vienna, Austria. Httpwww R-Proj Org [Internet]. 2016 [cited 2025 Sept 8]; Available from: https://cir.nii.ac.jp/crid/1574231874043578752

30. Sutter RW, Hardy IR, Kozlova IA, Tchoudnaia LM, Gluskevich TG, Marievsky V, et al. Immunogenicity of tetanus-diphtheria toxoids (Td) among Ukrainian adults: implications for diphtheria control in the Newly Independent States of the Former Soviet Union. J Infect Dis. 2000;181(Supplement_1):S197–202.

31. Frost WH. Infection, immunity and disease in the epidemiology of diphtheria, with special reference to some studies in Baltimore. J Prev Med. 1928;2(4):325–43.

32. Adetifa IMO, Karia B, Mutuku A, Bwanaali T, Makumi A, Wafula J, et al. Coverage and timeliness of vaccination and the validity of routine estimates: Insights from a vaccine registry in Kenya. Vaccine. 2018 Dec 18;36(52):7965–74.

33. Cutts FT, Izurieta HS, Rhoda DA. Measuring coverage in MNCH: design, implementation, and interpretation challenges associated with tracking vaccination coverage using household surveys. PLoS Med. 2013;10(5):e1001404.

34. WHO. Pertussis vaccines: WHO position paper—August 2015. Wkly Epidemiol Rec. 2015;90:433–58.

35. Chen Z, He Q. Immune persistence after pertussis vaccination. Hum Vaccines Immunother. 2017 Apr 3;13(4):744–56.

36. Immunization Data [Internet]. [cited 2025 Sept 10]. WHO Immunization Data portal - Detail Page. Available from: https://immunizationdata.who.int/global/wiise-detail-page

37. Daily Nation [Internet]. 2022 [cited 2025 Sept 10]. IMMUNISATION WEEK: Tetanus, diphtheria vaccinations provide lifetime immunity for your baby. Available from: https://nation.africa/kenya/health/tips/immunisation-week-tetanus-diphtheria-vaccinations-provide-lifetime-immunity-for-your-baby-3794648

38. Geteri F, Dawa J, Gachohi J, Kadivane S, Humwa F, Okunga E. A recent history of disease outbreaks in Kenya, 2007–2022: Findings from routine surveillance data. BMC Res Notes. 2024 Oct 15;17:309.

39. Khetsuriani N, Zakikhany K, Jabirov S, Saparova N, Ursu P, Wannemuehler K, et al. Seroepidemiology of diphtheria and tetanus among children and young adults in Tajikistan: nationwide population-based survey, 2010. Vaccine. 2013;31(42):4917–22.

40. McQuillan GM, Kruszon-Moran D, Deforest A, Chu SY, Wharton M. Serologic Immunity to Diphtheria and Tetanus in the United States. Ann Intern Med. 2002 May 7;136(9):660–6.

41. Crossley K, Irvine P, Warren JB, Lee BK, Mead K. Tetanus and diphtheria immunity in urban Minnesota adults. Jama. 1979;242(21):2298–300.

42. Kimura M, Kuno-Sakai H, Sato Y, Kamiya H, Nii R, Isomura S, et al. A comparative trial of the reactogenicity and immunogenicity of Takeda acellular pertussis vaccine combined with tetanus and diphtheria toxoids: outcome in 3-to 8-month-old infants, 9-to 23-month-old infants and children, and 24-to 30-month-old children. Am J Dis Child. 1991;145(7):729–33.

43. Hammarlund E, Thomas A, Poore EA, Amanna IJ, Rynko AE, Mori M, et al. Durability of vaccine-induced immunity against tetanus and diphtheria toxins: a cross-sectional analysis. Clin Infect Dis. 2016;62(9):1111–8.

44. Borrow R, Balmer P, Roper MH. Immunological basis for immunization: Molule 3 : Tetanus (update 2006). Geneva: World Health Organization; 2007.

45. Scobie HM, Patel M, Martin D, Mkocha H, Njenga SM, Odiere MR, et al. Tetanus Immunity Gaps in Children 5–14 Years and Men ≥ 15 Years of Age Revealed by Integrated Disease Serosurveillance in Kenya, Tanzania, and Mozambique. Am J Trop Med Hyg. 2017 Feb 8;96(2):415–20.

46. Galazka A, Dittmann S. The changing epidemiology of diphtheria in the vaccine era. J Infect Dis. 2000;181(Supplement_1):S2–9.

47. Edmunds WJ, Pebody RG, Aggerback H, Baron S, Berbers G, Conyn-Van Spaendonck MAE, et al. The sero-epidemiology of diphtheria in Western Europe. Epidemiol Infect. 2000;125(1):113–25.

48. Galazka AM, Robertson SE. Diphtheria: Changing patterns in the developing world and the industrialized world. Eur J Epidemiol. 1995 Feb;11(1):107–17.

49. Gaayeb L, Sarr JB, Ndiath MO, Hanon JB, Debrie AS, Seck M, et al. Seroprevalence of pertussis in Senegal: a prospective study. PLoS One. 2012;7(10):e48684.

50. Plotkin SA. Correlates of Protection Induced by Vaccination. Clin Vaccine Immunol. 2010 July;17(7):1055–65.

51. Wendelboe AM, Van Rie A, Salmaso S, Englund JA. Duration of immunity against pertussis after natural infection or vaccination. Pediatr Infect Dis J. 2005 May;24(5 Suppl):S58–61.

52. Chen Z, He Q. Immune persistence after pertussis vaccination. Hum Vaccines Immunother. 2017 Jan 3;13(4):744–56.

53. Sheng Y, Ma S, Zhou Q, Xu J. Pertussis resurgence: epidemiological trends, pathogenic mechanisms, and preventive strategies. Front Immunol [Internet]. 2025 July 10 [cited 2025 Sept 10];16. Available from: https://www.frontiersin.org/journals/immunology/articles/10.3389/fimmu.2025.1618883/full

