## Supplemental files for "Seroprevalence of antibodies against Diphtheria, Tetanus and Pertussis over a 12-year period in children in Kilifi, Kenya (2009-2021)"

Table s1: Standardized age-specific diphtheria and tetanus seroprevalence estimates categorised into minimal seroprotection (0.011≤IgG<0.1 IU/ml), full seroprotection (0.1≤IgG<1 IU/ml) and long-term protection (IgG≥1 IU/ml). Seroprevalence estimates were also adjusted for test performance using Bayesian modelling.

| Survey year | Age in years |  | diphtheria | | | tetanus | | |
| --- | --- | --- | --- | --- | --- | --- | --- | --- |
|  | Level of seroprotection | n | Minimal% | Full% | Long-term% | Minimal% | Full% | Long-term% |
| 2009 | <1yr | 14 | 40[20-63] | 46[18-72] | 02[00-07] | 10[02-27] | 49[23-73] | 25[08-50] |
|  | 1-4yrs | 133 | 56[47-65] | 08[01-19] | 01[00-02] | 21[14-29] | 60[46-70] | 05[01-11] |
|  | 5-9yrs | 202 | 50[41-59] | 08[01-17] | 01[00-03] | 32[25-39] | 30[11-43] | 21[14-28] |
|  | 10-14yrs | 16 | 45[26-68] | 10[01-30] | 01[00-08] | 16[04-36] | 23[05-46] | 49[26-72] |
|  | Total | 365 | 50[41-58] | 11[05-22] | 01[00-04] | 23[17-30] | 37[24-49] | 25[17-33] |
|  | P value |  | 0.054 | 0.001 | 0.483 | 0.038 | <0.001 | <0.001 |
| 2011 | <1yr | 14 | 21[06-39] | 72[44-91] | 03[00-14] | 10[02-27] | 19[04-43] | 67[43-88] |
|  | 1-4yrs | 82 | 13[05-24] | 59[46-69] | 10[04-17] | 05[01-12] | 26[07-42] | 57[46-68] |
|  | 5-9yrs | 142 | 51[42-60] | 09[01-20] | 01[00-04] | 38[31-47] | 23[07-37] | 18[10-25] |
|  | 10-14yrs | 70 | 57[44-69] | 10[01-23] | 01[00-05] | 21[12-31] | 24[06-42] | 40[28-54] |
|  | Total | 308 | 40[34-46] | 28[21-36] | 04[02-07] | 22[17-27] | 24[09-37] | 39[33-45] |
|  | P value |  | <0.001 | <0.001 | 0.001 | <0.001 | 0.444 | <0.001 |
| 2013 | <1yr | 7 | 34[13-62] | 52[17-83] | 02[00-09] | 09[01-29] | 28[06-59] | 55[24-82] |
|  | 1-4yrs | 123 | 50[40-59] | 37[24-47] | 01[00-04] | 09[03-16] | 57[43-69] | 23[14-31] |
|  | 5-9yrs | 148 | 51[42-60] | 15[05-26] | 02[00-04] | 30[22-39] | 40[23-53] | 12[06-19] |
|  | 10-14yrs | 128 | 58[48-67] | 11[02-22] | 01[00-02] | 28[20-37] | 20[04-36] | 31[22-39] |
|  | Total | 406 | 52[45-58] | 22[15-31] | 01[00-03] | 22[17-27] | 38[25-49] | 24[18-29] |
|  | P value |  | 0.256 | <0.001 | 0.384 | <0.001 | <0.001 | <0.001 |
| 2015 | <1yr | 35 | 19[08-34] | 68[50-82] | 07[01-16] | 04[01-13] | 38[15-57] | 48[32-64] |
|  | 1-4yrs | 144 | 53[43-62] | 28[17-38] | 01[00-03] | 11[04-17] | 64[51-73] | 15[09-23] |
|  | 5-9yrs | 153 | 49[40-58] | 15[04-26] | 01[00-02] | 36[29-45] | 39[23-52] | 08[02-14] |
|  | 10-14yrs | 44 | 59[43-73] | 07[01-20] | 01[00-04] | 33[20-48] | 21[04-40] | 22[11-37] |
|  | Total | 376 | 51[44-58] | 20[13-28] | 01[01-03] | 26[20-32] | 41[28-51] | 17[11-23] |
|  | P value |  | <0.001 | <0.001 | <0.001 | <0.001 | <0.001 | <0.001 |
| 2017 | <1yr | 27 | 18[06-35] | 60[38-78] | 11[03-25] | 08[02-22] | 45[20-65] | 32[18-52] |
|  | 1-4yrs | 160 | 48[38-57] | 34[24-44] | 01[00-04] | 08[03-13] | 69[59-78] | 15[08-21] |
|  | 5-9yrs | 185 | 51[42-59] | 17[07-27] | 02[0-05] | 40[33-47] | 40[25-52] | 04[01-09] |
|  | 10-14yrs | 49 | 45[31-59] | 08[01-22] | 02[00-06] | 53[39-66] | 18[04-35] | 08[01-18] |
|  | Total | 421 | 46[39-53] | 22[15-30] | 03[01-04] | 33[27-38] | 42[31-52] | 10[06-15] |
|  | P value |  | <0.001 | <0.001 | <0.001 | <0.001 | <0.001 | <0.001 |
| 2019 | <1yr | 39 | 25[12-40] | 57[39-74] | 08[02-20] | 05[01-14] | 29[08-48] | 55[40-71] |
|  | 1-4yrs | 205 | 52[43-60] | 33[23-42] | 02[00-04] | 10[05-15] | 71[62-80] | 08[03-14] |
|  | 5-9yrs | 220 | 51[44-58] | 10[02-20] | 02[00-04] | 33[27-40] | 45[30-56] | 04[01-09] |
|  | 10-14yrs | 56 | 55[41-68] | 08[01-21] | 04[00-09] | 37[24-50] | 30[10-47] | 09[02-19] |
|  | Total | 520 | 51[44-57] | 19[13-27] | 03[01-05] | 26[20-31] | 46[34-56] | 11[06-15] |
|  | P value |  | 0.003 | <0.001 | 0.002 | <0.001 | <0.001 | <0.001 |
| 2021 | <1yr | 8 | 27[10-50] | 35[08-68] | 07[01-29] | 09[01-30] | 27[06-56] | 59[28-85] |
|  | 1-4yrs | 85 | 30[18-41] | 51[39-63] | 04[01-09] | 07[02-14] | 55[37-68] | 25[15-35] |
|  | 5-9yrs | 102 | 52[41-62] | 29[17-42] | 03[00-07] | 24[15-33] | 49[34-63] | 11[04-20] |
|  | 10-14yrs | 95 | 44[32-55] | 24[11-37] | 05[01-10] | 38[28-49] | 21[05-37] | 19[11-28] |
|  | Total | 290 | 41[34-48] | 34[26-43] | 04[02-07] | 23[17-28] | 41[29-52] | 21[15-26] |
|  | P value |  | 0.004 | 0.004 | 0.109 | <0.001 | <0.001 | <0.001 |

Table s2. Standardized Geometric Mean concentrations (GMCs) for diphtheria and tetanus by age group and year using a reference population to facilitate comparison across years.

| Survey year | 2009 | | | 2011 | | | 2013 | | | 2015 | | | 2017 | | | 2019 | | | 2021 | | |
| --- | --- | --- | --- | --- | --- | --- | --- | --- | --- | --- | --- | --- | --- | --- | --- | --- | --- | --- | --- | --- | --- |
| Diphtheria | N | GMC [95% CI] | | n | GMC [95% CI] | | n | GMC [95% CI] | | n | GMC [95% CI] | | n | GMC [95% CI] | | n | GMC [95% CI] | | n | GMC [95% CI] | |
| Age categories |  |  |  |  |  |  |  |  |  |  |  |  |  |  |  |  |  |  |  |  |  |
| <1yr | 14 | 0.15 | [0.08-0.27] | 14 | 0.31 | [0.16-0.59] | 7 | 0.22 | [0.08-0.63] | 35 | 0.29 | [0.20-0.43] | 27 | 0.29 | [0.15-0.56] | 39 | 0.27 | [0.19-0.38] | 8 | 0.26 | [0.05-1.29] |
| 1-4yrs | 133 | 0.03 | [0.02-0.05] | 82 | 0.18 | [0.09-0.39] | 123 | 0.11 | [0.07-0.17] | 144 | 0.06 | [0.04-0.10] | 160 | 0.09 | [0.06-0.12] | 205 | 0.08 | [0.06-0.11] | 85 | 0.13 | [0.07-0.26] |
| 5-9yrs | 202 | 0.02 | [0.01-0.04] | 142 | 0.03 | [0.01-0.06] | 148 | 0.04 | [0.02-0.08] | 153 | 0.03 | [0.02-0.05] | 185 | 0.04 | [0.02-0.07] | 220 | 0.04 | [0.02-0.07] | 102 | 0.09 | [0.05-0.16] |
| 10-14yrs | 16 | 0.01 | [0.00-0.01] | 70 | 0.02 | [0.01-0.03] | 128 | 0.03 | [0.02-0.05] | 44 | 0.03 | [0.01-0.09] | 49 | 0.02 | [0.01-0.08] | 56 | 0.04 | [0.01-0.13] | 95 | 0.06 | [0.03-0.13] |
| Total | 365 | 0.03 | [0.02-0.05] | 308 | 0.09 | [0.04-0.18] | 406 | 0.07 | [0.04-0.13] | 376 | 0.06 | [0.04-0.10] | 421 | 0.07 | [0.04-0.12] | 520 | 0.07 | [0.04-0.12] | 290 | 0.10 | [0.05-0.26] |
| P_value |  | <0.001 | |  | <0.001 | |  | <0.001 | |  | <0.001 | |  | <0.001 | |  | <0.001 | |  | <0.001 | |
| Tetanus |  |  |  |  |  |  |  |  |  |  |  |  |  |  |  |  |  |  |  |  |  |
| <1yr | 14 | 0.54 | [0.30-0.98] | 14 | 1.25 | [0.65-2.42] | 7 | 1.67 | [0.53-5.27] | 35 | 1.03 | [0.76-1.39] | 27 | 0.82 | [0.54-1.25] | 39 | 1.03 | [0.75-1.41] | 8 | 1.20 | [0.44-3.23] |
| 1-4yrs | 133 | 0.22 | [0.15-0.33] | 82 | 1.06 | [0.58-1.96] | 123 | 0.58 | [0.40-0.85] | 144 | 0.42 | [0.30-0.60] | 160 | 0.37 | [0.27-0.52] | 205 | 0.30 | [0.22-0.41] | 85 | 0.49 | [0.29-0.86] |
| 5-9yrs | 202 | 0.27 | [0.15-0.50] | 142 | 0.18 | [0.09-0.37] | 148 | 0.17 | [0.10-0.30] | 153 | 0.15 | [0.09-0.25] | 185 | 0.11 | [0.07-0.18] | 220 | 0.13 | [0.08-0.21] | 102 | 0.25 | [0.15-0.44] |
| 10-14yrs | 16 | 0.09 | [0.04-0.22] | 70 | 0.31 | [0.16-0.62] | 128 | 0.37 | [0.15-0.93] | 44 | 0.41 | [0.14-1.63] | 49 | 0.09 | [0.02-1.81] | 56 | 0.10 | [0.04-0.31] | 95 | 0.16 | [0.06-0.42] |
| Total | 365 | 0.22 | [0.13-0.40] | 308 | 0.55 | [0.29-1.04] | 406 | 0.46 | [0.23-1.00] | 376 | 0.37 | [0.21-0.85] | 421 | 0.23 | [0.15-0.85] | 520 | 0.23 | [0.16-0.38] | 290 | 0.36 | [0.18-0.75] |
| P_value |  | 0.012 | |  | <0.001 | |  | <0.001 | |  | <0.001 | |  | <0.001 | |  | <0.001 | |  | <0.001 | |

Table s3. Standardized age-specific diphtheria and tetanus seroprevalence estimates and Geometric Mean concentrations (GMCs) in adults. Seroprevalence estimates were also adjusted for test performance using Bayesian modelling

|  | | Level of seroprotection | | | | | |  | |
| --- | --- | --- | --- | --- | --- | --- | --- | --- | --- |
| Diphtheria | n | Minimal% [95% CI] | | Full% [95% CI] | | Long-term% [95% CI] | | GMC [95% CI] | |
| 15_19 | 53 | 52 | [39-65] | 13 | [02-29] | 01 | [00-04] | 0.03 | [0.01-0.09] |
| 20_24 | 51 | 41 | [27-55] | 21 | [06-38] | 01 | [00-04] | 0.04 | [0.01-0.19] |
| 25_29 | 49 | 44 | [31-57] | 15 | [02-31] | 01 | [00-04] | 0.03 | [0.01-0.11] |
| 30_34 | 51 | 54 | [40-67] | 07 | [01-22] | 01 | [00-04] | 0.03 | [0.01-0.08] |
| 35_39 | 49 | 51 | [37-65] | 11 | [01-26] | 01 | [00-04] | 0.04 | [0.01-0.14] |
| 40_44 | 50 | 59 | [45-72] | 08 | [01-21] | 01 | [00-04] | 0.03 | [0.01-0.10] |
| 45-49 | 50 | 57 | [43-70] | 08 | [01-22] | 01 | [00-06] | 0.03 | [0.01-0.13] |
| 50_54 | 48 | 41 | [27-55] | 22 | [05-39] | 04 | [01-10] | 0.06 | [0.02-0.75] |
| 55_59 | 50 | 43 | [30-57] | 12 | [02-28] | 01 | [00-06] | 0.03 | [0.01-0.16] |
| 60_64 | 49 | 58 | [44-70] | 09 | [01-25] | 02 | [00-06] | 0.04 | [0.01-0.20] |
| 65+ | 50 | 62 | [48-74] | 10 | [01-25] | 01 | [00-04] | 0.02 | [0.00-0.90] |
| Total | 550 | 50 | [45-54] | 17 | [14-21] | 01 | [00-02] | 0.04 | [0.01-0.24] |
| P value |  | 0.10 | | 0.53 | | 0.11 | | 0.76 | |
| Tetanus |  |  |  |  |  |  |  |  |  |
| 15_19 | 53 | 42 | [30-56] | 19 | [03-37] | 10 | [02-21] | 0.12 | [0.00-1.76] |
| 20_24 | 51 | 15 | [06-26] | 13 | [02-31] | 56 | [42-70] | 0.88 | [0.01-4.19] |
| 25_29 | 49 | 03 | [01-09] | 19 | [04-39] | 60 | [46-74] | 1.70 | [0.02-4.02] |
| 30_34 | 51 | 05 | [01-13] | 10 | [02-25] | 70 | [56-82] | 1.47 | [0.02-3.92] |
| 35_39 | 49 | 08 | [02-18] | 23 | [05-42] | 48 | [35-63] | 1.42 | [0.02-4.29] |
| 40_44 | 50 | 08 | [02-17] | 30 | [09-49] | 37 | [23-52] | 0.67 | [0.01-2.65] |
| 45-49 | 50 | 20 | [10-33] | 26 | [06-45] | 28 | [15-42] | 0.32 | [0.00-1.98] |
| 50_54 | 48 | 10 | [03-21] | 28 | [07-47] | 41 | [27-54] | 1.01 | [0.03-3.89] |
| 55_59 | 50 | 18 | [09-30] | 26 | [05-44] | 31 | [19-47] | 0.45 | [0.01-1.23] |
| 60_64 | 49 | 17 | [08-29] | 27 | [08-46] | 24 | [14-38] | 0.29 | [0.01-1.41] |
| 65+ | 50 | 26 | [15-39] | 22 | [05-42] | 20 | [09-33] | 0.06 | [0.00-1.12] |
| Total | 550 | 09 | [06-13] | 29 | [25-34] | 36 | [32-41] | 0.76 | [0.01-2.87] |
| P value |  | <0.001 | | 0.19 | | <0.001 | | <0.001 | |

Table s4. Standardized age-specific pertussis seroprevalence estimates and Geometric Mean concentrations (GMCs) in children.; IgG ≥125 EU/mL (infection in the past 6 months), 62.5≤IgG <125 EU/mL (infection in the past 12 months), 20≤IgG <62.5 EU/mL (infection in the past ≥12 months or vaccination response) and 0≤IgG <20 EU/mL (no recent infection). Seroprevalence estimates were also adjusted for test performance using Bayesian modelling.

|  |  |  | Level of seroprotection | | | | GMCs [95% CI] | |
| --- | --- | --- | --- | --- | --- | --- | --- | --- |
| Survey year | Age category | n | 0≤IgG <20 EU/mL | 20≤IgG <62.5 EU/mL | 62.5≤IgG <125 EU/mL | IgG≥125 EU/mL |  | |
| 2009 | <1yr | 14 | 63[26-81] | 10[02-24] | 05[00-39] | 00[00-18] | 4.04 | [1.48-11.00] |
|  | 1-4yrs | 133 | 66[33-77] | 11[02-22] | 00[00-02] | 00[00-04] | 5.02 | [2.40-10.50] |
|  | 5-9yrs | 202 | 61[24-72] | 08[01-18] | 04[00-09] | 02[02-07] | 7.49 | [3.83-14.83] |
|  | 10-14yrs | 16 | 55[17-72] | 16[03-32] | 00[00-15] | 03[03-21] | 1.82 | [0.62-5.41] |
|  | Total | 365 | 61[28-71] | 11[03-21] | 01[00-05] | 01[00-05] | 4.81 | [2.28-10.45] |
|  | P value |  | 0.25 | 0.11 | 0.07 | 0.52 | 0.12 | |
| 2011 | <1yr | 14 | 58[21-75] | 15[3-31] | 01[00-27] | 00[00-17] | 7.23 | [2.87-18.20] |
|  | 1-4yrs | 82 | 74[51-84] | 08[01-19] | 00[00-03] | 00[00-01] | 3.24 | [1.56-6.84] |
|  | 5-9yrs | 142 | 59[21-71] | 16[04-28] | 00[00-04] | 00[00-02] | 9.37 | [5.08-17.56] |
|  | 10-14yrs | 70 | 46[09-61] | 22[07-34] | 09[02-19] | 00[00-03] | 9.23 | [4.33-19.95] |
|  | Total | 308 | 59[29-69] | 15[06-25] | 01[00-05] | 00[00-03] | 7.44 | [3.70-15.30] |
|  | P value |  | <0.001 | 0.02 | 0.01 | 0.33 | <0.001 | |
| 2013 | <1yr | 7 | 52[17-71] | 17[03-36] | 14[02-62] | 00[00-32] | 34.80 | [14.00-86.30] |
|  | 1-4yrs | 123 | 58[23-71] | 16[04-27] | 03[00-09] | 00[00-01] | 7.88 | [4.27-14.83] |
|  | 5-9yrs | 148 | 67[36-78] | 08[01-19] | 00[00-03] | 01[01-07] | 5.45 | [2.44-12.40] |
|  | 10-14yrs | 128 | 55[17-67] | 18[06-29] | 00[00-04] | 01[01-07] | 11.61 | [4.82-28.79] |
|  | Total | 406 | 60[27-69] | 14[05-23] | 00[00-03] | 00[00-04] | 10.11 | [4.51-23.36] |
|  | P value |  | 0.02 | 0.02 | 0.03 | 0.08 | 0.01 | |
| 2015 | <1yr | 35 | 58[19-73] | 13[02-27] | 06[00-21] | 00[00-04] | 5.22 | [2.68-10.10] |
|  | 1-4yrs | 144 | 56[18-68] | 19[07-31] | 00[00-05] | 00[00-02] | 9.75 | [5.58-17.09] |
|  | 5-9yrs | 153 | 68[40-79] | 09[01-20] | 00[00-04] | 00[00-01] | 4.73 | [2.35-9.61] |
|  | 10-14yrs | 44 | 59[21-73] | 11[02-23] | 03[00-15] | 03[02-15] | 7.63 | [2.98-24.81] |
|  | Total | 376 | 61[30-71] | 13[05-23] | 00[00-04] | 00[00-04] | 7.07 | [3.48-16.41] |
|  | P value |  | 0.08 | 0.07 | 0.51 | 0.16 | <0.001 | |
| 2017 | <1yr | 27 | 60[23-75] | 11[02-24] | 06[00-26] | 02[02-20] | 8.65 | [3.81-19.70] |
|  | 1-4yrs | 160 | 58[22-70] | 18[07-30] | 00[00-02] | 00[00-01] | 7.83 | [4.39-14.07] |
|  | 5-9yrs | 185 | 71[46-80] | 09[02-20] | 00[00-01] | 00[00-00] | 4.23 | [2.09-8.80] |
|  | 10-14yrs | 49 | 59[23-73] | 12[02-24] | 05[00-17] | 00[00-07] | 5.49 | [1.77-22.01] |
|  | Total | 421 | 63[34-73] | 12[05-23] | 00[00-01] | 00[00-05] | 5.95 | [2.77-15.10] |
|  | P value |  | 0.04 | 0.06 | 0.02 | 0.18 | 0.004 | |
| 2019 | <1yr | 39 | 64[28-78] | 10[02-22] | 00[00-12] | 02[02-15] | 4.14 | [2.03-8.47] |
|  | 1-4yrs | 205 | 66[36-76] | 13[03-24] | 00[00-00] | 00[00-00] | 5.56 | [3.16-9.78] |
|  | 5-9yrs | 220 | 59[26-70] | 14[04-24] | 00[00-02] | 00[00-05] | 7.58 | [3.96-14.92] |
|  | 10-14yrs | 56 | 56[16-71] | 14[03-27] | 01[00-12] | 03[03-14] | 11.18 | [5.53-26.00] |
|  | Total | 520 | 61[28-70] | 13[04-24] | 00[00-01] | 00[00-05] | 7.86 | [4.08-16.39] |
|  | P value |  | 0.23 | 0.53 | 0.54 | 0.07 | 0.003 | |
| 2021 | <1yr | 8 | 58[19-76] | 12[02-29] | 07[00-44] | 07[07-44] | 14.90 | [3.11-71.20] |
|  | 1-4yrs | 85 | 45[09-60] | 25[11-37] | 08[02-17] | 00[00-02] | 13.02 | [5.82-29.19] |
|  | 5-9yrs | 102 | 58[21-70] | 15[04-27] | 04[00-12] | 00[00-00] | 8.27 | [3.55-19.30] |
|  | 10-14yrs | 95 | 50[13-66] | 17[05-30] | 07[01-16] | 00[00-05] | 12.82 | [5.02-34.04] |
|  | Total | 290 | 49[42-55] | 21[16-27] | 06[03-11] | 00[00-05] | 11.48 | [4.61-30.30] |
|  | P value |  | 0.11 | 0.22 | 0.91 | 0.06 | 0.23 | |

Table s5. Standardized age-specific pertussis seroprevalence estimates and Geometric Mean concentrations (GMCs) in adults. Seroprevalence estimates were also adjusted for test performance using Bayesian modelling.

|  |  |  | Level of seroprotection | | | | GMCs | |
| --- | --- | --- | --- | --- | --- | --- | --- | --- |
| Survey year | Age category | n | 0≤IgG <20 EU/mL | 20≤IgG <62.5 EU/mL | 62.5≤IgG <125 EU/mL | IgG≥125 EU/mL |  |  |
| 2021 | 15-19yrs | 53 | 58[20-73] | 15[03-30] | 00[00-07] | 00[00-07] | 15.51 | [6.90-35.41] |
|  | 20-24yrs | 51 | 55[17-71] | 16[03-29] | 02[00-13] | 02[00-13] | 15.84 | [4.97-40.90] |
|  | 25-29yrs | 49 | 59[23-74] | 14[03-28] | 02[00-14] | 02[00-14] | 08.46 | [2.90-26.54] |
|  | 30-34yrs | 51 | 63[29-76] | 11[02-23] | 04[00-16] | 04[00-16] | 11.75 | [5.06-29.02] |
|  | 35-39yrs | 49 | 58[18-71] | 18[04-32] | 00[00-11] | 00[00-11] | 12.87 | [4.34-42.40] |
|  | 40-44yrs | 50 | 63[27-76] | 14[03-28] | 00[00-05] | 00[00-5] | 11.05 | [4.47-28.07] |
|  | 45-49yrs | 50 | 58[21-73] | 15[03-28] | 00[00-11] | 00[00-11] | 11.82 | [4.64-42.87] |
|  | 50-54yrs | 48 | 65[28-78] | 11[02-24] | 00[00-12] | 00[00-12] | 15.26 | [5.31-52.50] |
|  | 55-59yrs | 50 | 62[25-76] | 15[03-28] | 00[00-05] | 00[00-05] | 09.32 | [3.28-33.62] |
|  | 60-64yrs | 49 | 53[15-69] | 19[05-33] | 05[00-17] | 05[00-17] | 16.15 | [8.04-35.59] |
|  | >=65yrs | 50 | 46[11-62] | 32[17-48] | 00[00-08] | 00[00-08] | 03.97 | [1.40-37.12] |
|  | Total | 550 | 63[58-67] | 23[19-27] | 00[00-02] | 00[00-01] | 11.88 | [4.71-35.81] |
|  | P value |  | 0.17 | 0.05 | 0.75 | 0.37 | 0.22 | |

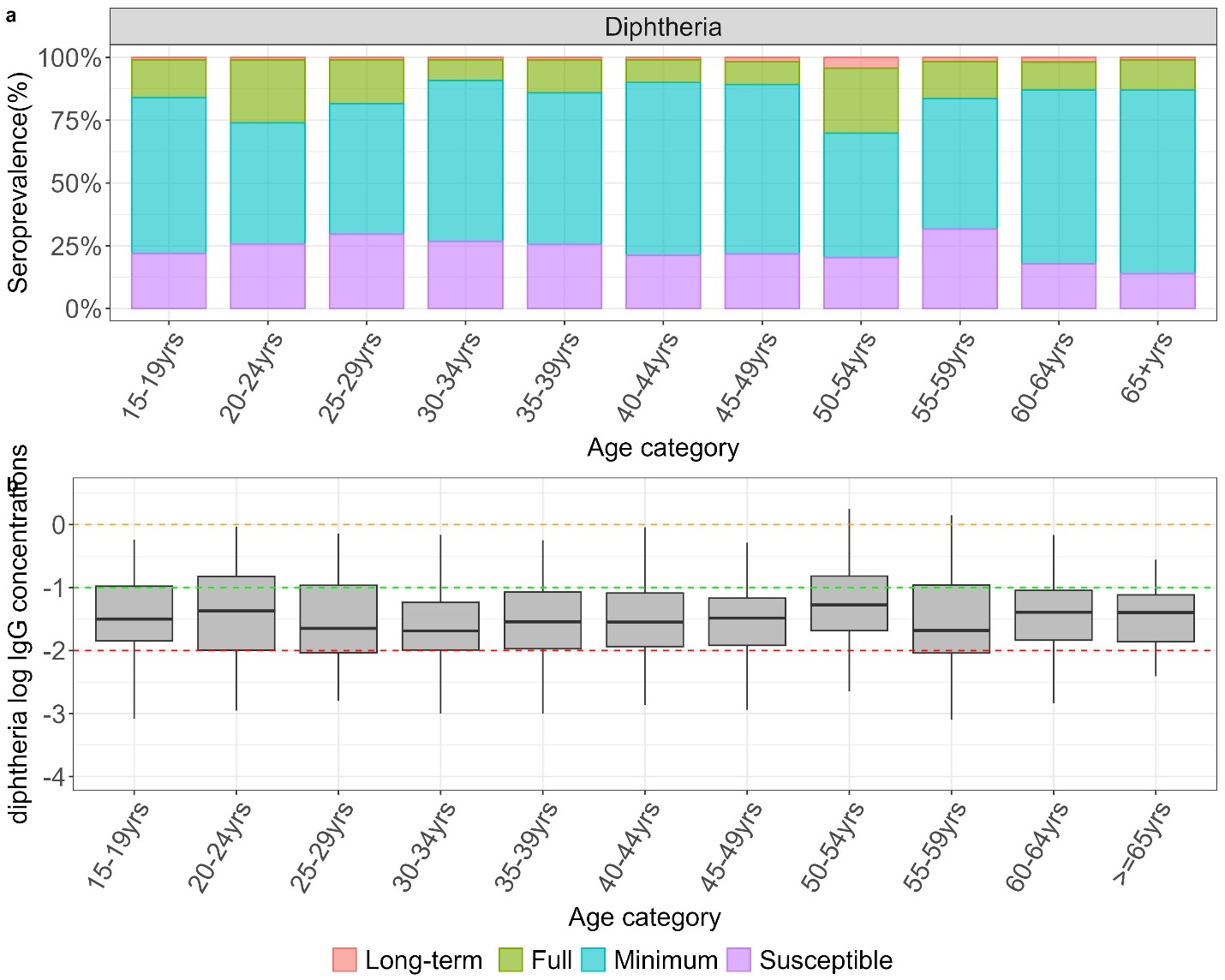

**Figure S1. a. Standardized diphtheria seroprevalence estimates in adults in 2021 adjusted for test performance using Bayesian multilevel regression and poststratification. b. Distribution of *log* IgG concentrations with median and IQR.**

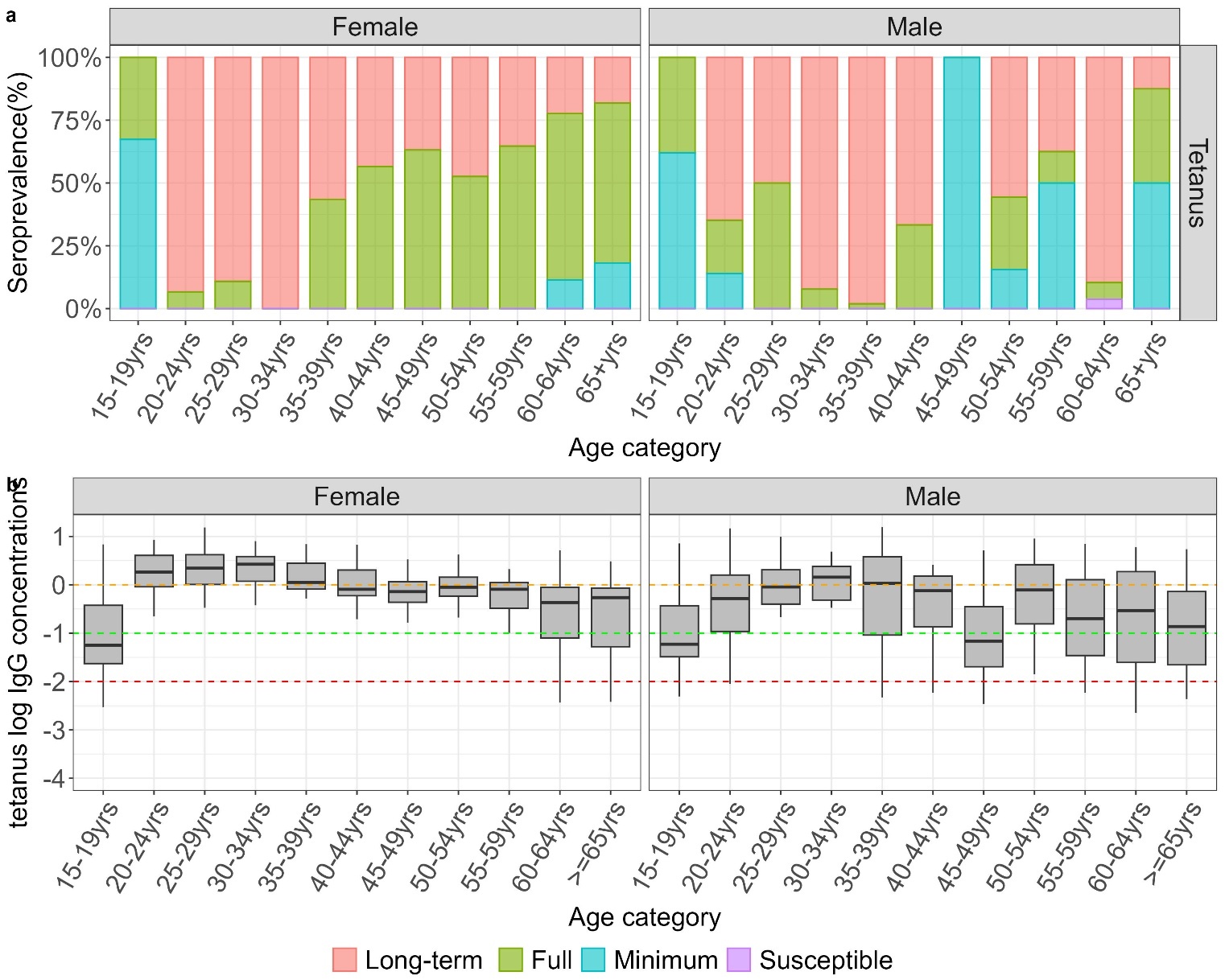

**Figure S2. a. Standardized tetanus seroprevalence estimates in adults in 2021 adjusted for test performance using Bayesian multilevel regression and poststratification. b. Distribution of *log* IgG concentrations with median and IQR.**

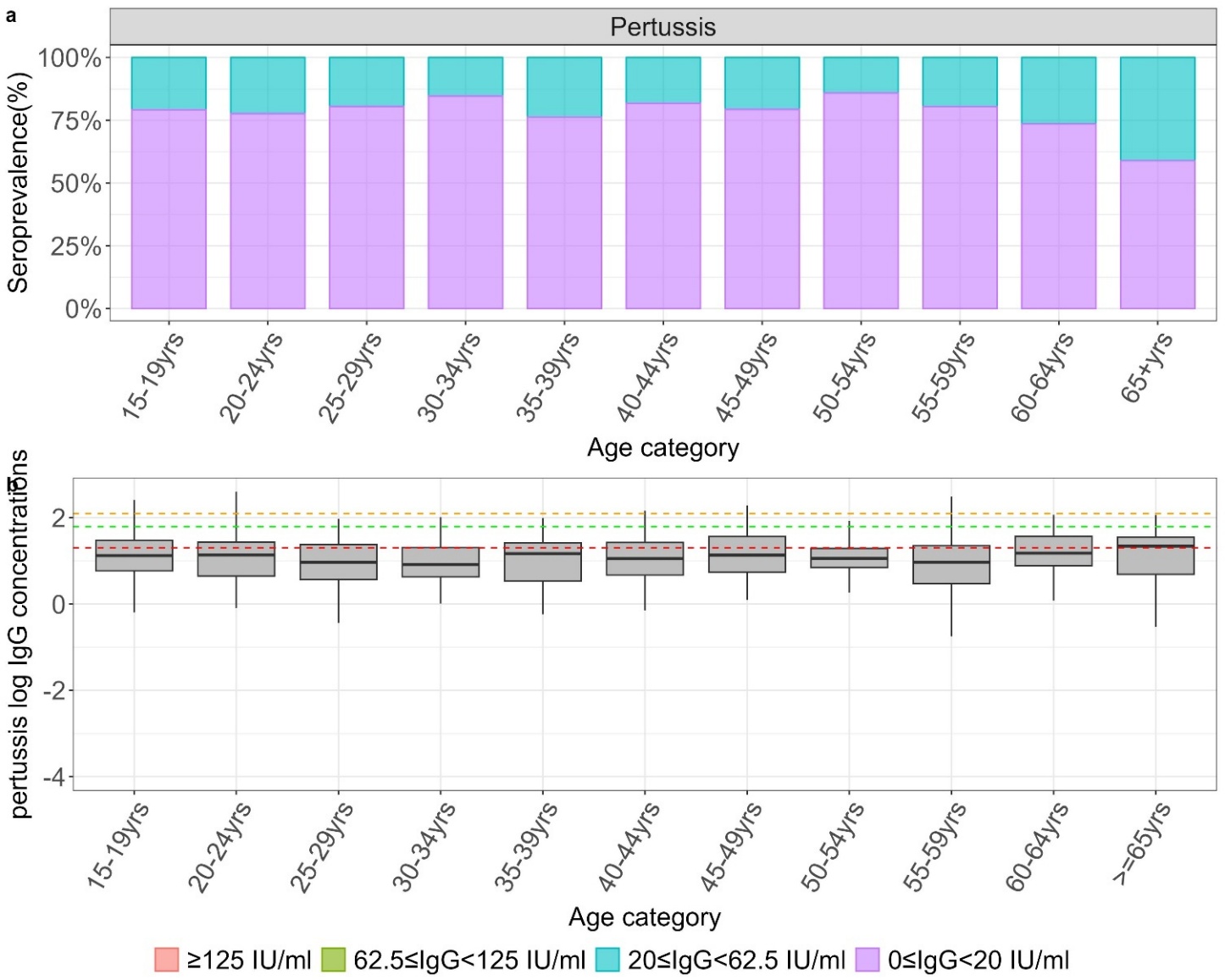

**Figure S3. a. Standardized pertussis seroprevalence estimates in adults in 2021 adjusted for test performance using Bayesian multilevel regression and poststratification. b. Distribution of log IgG concentrations with median and IQR.**
